# Immunogenicity and reactogenicity of mRNA COVID-19 vaccine booster administered by intradermal or intramuscular route in Thai Older adults

**DOI:** 10.1101/2022.12.16.22283601

**Authors:** Prasert Assantachai, Suvimol Niyomnaitham, Wichai Chatthanawaree, Somboon Intalapaporn, Weerasak Muangpaisan, Harisd Phannarus, Rangsimatiti Binda Saichompoo, Unchana Sura-amonrattana, Patimaporn Wongprompitak, Zheng Quan Toh, Paul V Licciardi, Kanjana Srisutthisamphan, Kulkanya Chokephaibulkit

## Abstract

**Introduction:** Intradermal (ID) vaccination may alleviate COVID-19 vaccine shortages and vaccine hesitancy due to systemic reactogenicity among older adults.

**Objectives:** To compare the immunogenicity and reactogenicity of fractional ID and standard intramuscular (IM) booster vaccination of mRNA-1273 and BNT162b2 vaccines in older adults.

**Methods:** Participants aged ≥65 years who previously vaccinated with 2-dose ChAdOx1 were randomized to receive one of the four booster vaccinations: 0.1mL ID mRNA-1273, 0.5mL IM mRNA-1273, 0.1mL ID BNT162b2 and 0.3mL IM BNT162b2. Immunogenicity as measured by anti-receptor binding domain (anti-RBD) IgG against Wuhan, neutralising antibody (NAb) against Wuhan and Omicron BA.1, BA.2 and BA.4/5, and IFNγ-producing cells. Local and systemic adverse effects (AEs) were self-reported via an electronic diary card.

**Results:** Of the 210 participants enrolled, 70.5% were female and median age was 77.5 years (interquartile range (IQR): 71.0-84.0). Following the booster dose, both ID vaccination induced 37% lower levels of anti-RBD IgG than IM vaccination of the same vaccine. NAb against ancestral and Omicron BA.1 strains was highest following IM mRNA-1273 (1,718 and 617), followed by ID mRNA-1273 (1,212 and 318), IM BNT162b2 (713 and 230), and ID BNT162b2 (587 and 148), respectively. Spike-specific IFNγ responses were similar or higher in the ID groups when compared with their respective IM groups. Vaccine delivery through ID route tended to have lower systemic AEs, although more local AEs reported in ID mRNA-1273 group.

**Conclusions:** Fractional ID vaccination induced immunogenicity and reactogenicity comparable to IM and may be an alternative option for older people.

**Key points:**

- Fractional dose intradermal mRNA COVID-19 booster vaccination induces robust immunogenicity in adults aged ≥65 years.
- For each vaccine, intradermal route induced lower humoral but similar or higher cellular immune responses than IM route.
- Intradermal mRNA-1273 vaccination induced similar immunogenicity to intramuscular BNT162b2 vaccination.
- Immune responses were marginally lower among participants aged ≥80 years than among participants aged 65-79 years.
- Systemic reaction was lower following intradermal mRNA COVID-19 vaccination compared with intramuscular vaccination.

## Introduction

COVID-19 booster vaccination has been found to improve protection against the SARS-CoV-2 variants, particularly against the omicron variants [1-4]. The booster vaccination has been recommended for older adults due to their aging immune system (immunosenecence) as well as the likelihood of having comorbidities that predisposes them to severe COVID-19. However, the vaccine coverage among older Thai adults, and in other settings, has been low, which could partly be attributed to vaccine hesitancy regarding adverse effects [2].

Intradermal (ID) vaccination of BCG and rabies vaccines have been given routinely in many settings. The dermis and epidermis layers are rich in antigen-presenting cells, and lower or fractional dosage of vaccine content are normally administered intradermally. ID administration for other vaccines such as inactivated polio, Hepatitis B and Influenza vaccines given at fractional dose is found to induce equivalent, or higher immunogenicity, and similar safety profiles compared to intramuscular (IM) or subcutaneous (SC) vaccination [5,6]. Hence, COVID-19 vaccination via the ID route may be considered as an alternative to IM vaccination. This would be particularly relevant in the context of limited COVID-19 vaccine supplies. Moreover, this alternative route using lower amount of vaccine may reduce vaccine hesitancy in some older people.

There are limited studies on COVID-19 vaccination given via the ID route. Most studies were conducted in healthy adults and found ID vaccination induce similar or slightly lower immune responses than IM, but with lower systemic adverse events [7]. Older adults have different immune composition under the skin and may respond to ID route differently. This study aims to compare the immunogenicity and reactogenicity of fractional dose mRNA-1273 (by Moderna) and BNT162b2 (by Pfizer) booster vaccination given via ID route with standard dose given mRNA-1273 and BNT162b2 via IM route in older adults previously vaccinated with two doses of ChAdOx1 (by Astra Zeneca).

## Methods

This open label study was conducted at single center, tertiary hospital in Bangkok, Thailand during the period of January to June 2022. Eligible subjects were individuals aged ≥65 years who have primarily received two doses of IM ChAdOx1 series 12-24 weeks earlier. Exclusion criteria included any previous infections of SARS-CoV-2, acute illness or inflammation, history of anaphylaxis to any vaccination or drugs, receipt of any vaccination within two previous weeks and receipt of any immunosuppressants or in the state of immunosuppression.

Written informed consent was obtained from the study subjects prior to any procedure. The subjects were randomized to one of the four vaccination groups: ID mRNA-1273 (20 mcg; 0.1 ml, n=35), IM mRNA-1273 (100 mcg; 0.5 ml, n=35), ID BNT162b2 (10 mcg; 0.1 ml, n=35 for each group of 65-<80 and ≥80 years of age) or IM BNT162b2 (30 mcg; 0.3 ml, n=35 for each group of 65-<80 and ≥80 years of age).

Blood samples were collected before and at 2-4 weeks after vaccination for immunogenicity evaluation of anti-SARS-CoV-2 receptor binding domain IgG (anti-RBD IgG) antibodies and pseudovirus neutralizing antibodies against ancestral Wuhan and Omicron variants (BA.1, BA.2, BA.4/5), as well as cellular immune response (by ELISpot) assessment. Another blood sample was collected at 12 weeks after vaccination to evaluate anti-RBD IgG levels. Baseline blood samples were tested for anti-nucleoprotein antibody (anti-NP) and anti-RBD IgG using qualitative assay (Abbott, List No. 06R86) on the ARCHITECT I System to exclude prior SARS-CoV-2 infection. Participants who were enrolled but later found to have positive anti-NP or anti-RBD at baseline were excluded from the analysis.

Participants were observed for ≥30 minutes following vaccination for any immediate adverse events. Participants and their caretakers (or person living together) were instructed to submit self-assessment report using an electronic diary (eDiary) in the Google Form for seven days for any adverse events (AEs). The solicited local AEs include pain, erythema, and swelling/induration at the injection site, and localized axillary lymphadenopathy or swelling or tenderness ipsilateral to the injection arm. The solicited systemic AEs included headache, fatigue, myalgia, arthralgia, diarrhea, dizziness, nausea/vomiting, rash, fever, and chills. The severity of solicited AEs was graded using a numerical scale from 1 to 4 based on the Common Terminology Criteria for Adverse Events – Version 5.0 guided by the United States National Cancer Institute (NCI/NIH) [8].

The study protocol was registered at Thai Clinical Trial Registry (TCTR20220112002) and was approved by the Siriraj Institutional Review Board (COA no. Si 001/2022).

### Measurement of SARS-CoV-2 anti-RBD

The anti-RBD IgG was measured using a chemiluminescent microparticle assay (SARS-CoV-2 IgG II Quant, Abbott, List No. 06S60). The level of antibodies quantified in arbitrary unit (AU/mL) and then converted into binding antibody unit per mL (BAU/mL) using the equation providedby the manufacturer (BAU/mL = 0.142 x AU/mL).

### Measurement of neutralising antibodies against SARS-CoV-2 variants

The pseudovirus neutralization test assay was carried out as previously published [9]. The neutralizing antibody titer (PVNT_50_) was defined as the highest test serum dilution that reduced the virus infectivity by 50% relative to the control wells with no serum. The minimum detection limit was 1:40; antibody titer of 1:20 was assigned to samples below the detection limit.

### Measurement of cellular immune response

Cellular immunity was determined by IFN-γ ELISpot (Mabtech, Nacka Strand, Sweden) to the ancestral strain on a subset of participants for each group (N=20). Peripheral blood mononuclear cells (PBMCs) were counted and stimulated with S-peptide consisting of 100 peptides from spike protein, and nucleoprotein-membrane protein-open reading frame proteins (NMO)-peptide pools consisting of 101 peptides from nucleocapsid (N), membrane (M), open reading frame (ORF) 1, non-structural protein (nsp) 3, ORF-3a, ORF-7a, and ORF8 proteins. Negative controls contained only cell culture media, while positive controls contained antiLJcluster of differentiation 3 (CD3) at a dilution of 1:1000. ELISpot plates were then incubated for 20 hours at 37°C and 5% CO_2_, washed and developed using a conjugated secondary antibody that bound to membrane-captured IFN-γ. The plates were read using IRIS (Mabtech) and spots were analyzed using Apex software 1.1 (Mabtech) and converted to spot-forming units (SFU) per million cells.

### Statistical Analysis

The anti-SARS-CoV-2 RBD IgG and PVNT_50_ titer were presented as GMC and geometric mean titers (GMT) with 95% confidence interval and were compared using unpaired t-test between the groups. The AEs endpoints were presented as frequencies and compared using Chi-square test between the groups. Using the results from our previous study [10], a sample size of 35 subjects per group provided 85% power to determine differences in antibody concentrations between ID and IM vaccination groups. GraphPad Prism 9 version 9.2.0 (283) (GraphPad Software, CA, USA) was used to perform all the statistical analyses except for the ANOVA analysis of the anti-RBD IgG among different age groups which was handled using STATA version 17 (StataCorp, LP, College Station, TX, USA).

## Results

### Participant’s baseline characteristics

A total of 231 participants were screened, and 211 were enrolled and randomized to one of the four vaccination groups; in BNT162b2 groups, participants were recruited into two age groups (Supplementary Figure S1). Among 210 participants included in the analysis, majority (70.5%) were female, median age was 77.5 years (interquartile range (IQR): 71.0-84.0) and median body mass index was 24.5 kg/m^2^ (IQR: 21.6-26.7 kg/m^2^). Majority (81.9%) of the participants have at least one health condition, and the median Charlson’s comorbidity index was 4.0 (IQR: 3.0-5.0), as shown in Table 1.

**Table 1.**
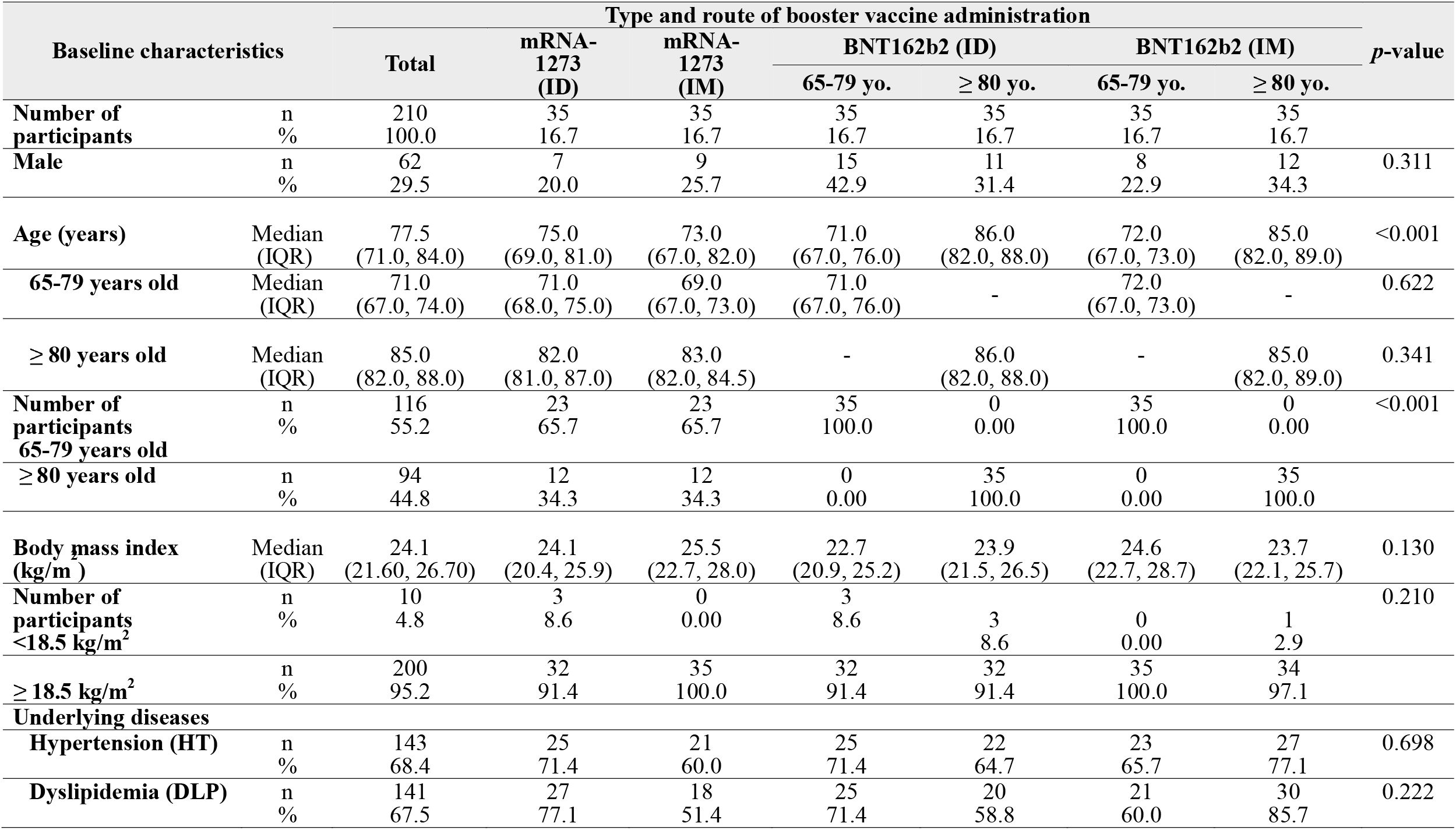
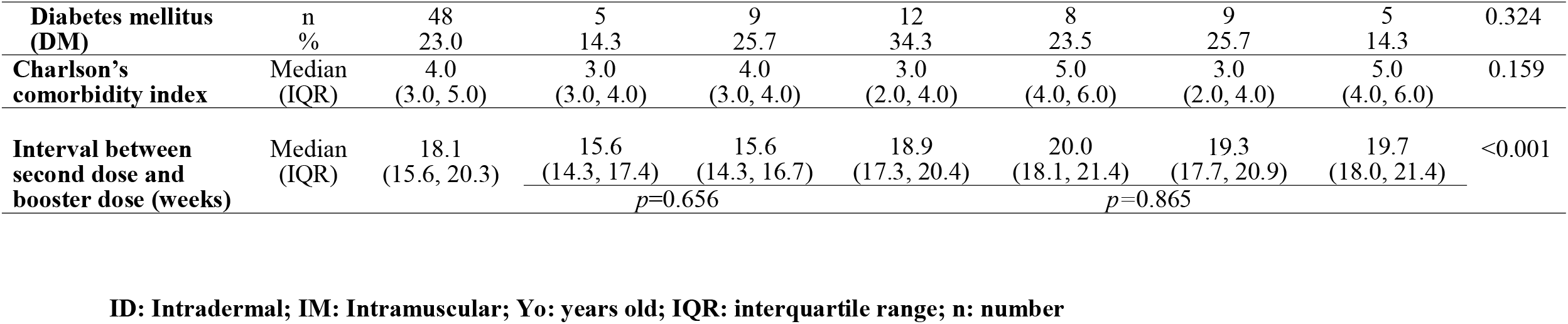
Baseline characteristics of the subjects by routes of administration and types of COVID-19 vaccines in booster dose

### Anti-SARS-CoV-2 RBD IgG responses

There were no statistical differences in anti-RBD IgG at baseline between the vaccine groups (Figure 1 and supplementary Table S1). At 2-4 weeks after booster vaccination, the GMC increased 29.2, 81.1, 35.3, and 53.3 times from baseline for ID mRNA-1273, IM mRNA-1273, ID BNT162b2, and IM BNT162b2 groups, respectively (*p*<0.001) (Figure 1 and Supplementary Table S1). Overall, the anti-RBD IgG GMC following mRNA-1273 vaccinations were higher than BNT162b2, regardless of administration routes. Participants who received IM mRNA-1273 had significantly higher anti-RBD IgG than IM BNT162b2 (GMC of IM mRNA-1273: 3,837.12 BAU/mL, GMC of IM BNT162b2: 2,530.58 BAU/mL, *p*=0.021). ID vaccination of either mRNA-1273 or BNT162b2 had 37% lower anti-RBD IgG responses than IM vaccination of the respective vaccine (*p*=0.028 for mRNA-1273 and *p*=0.005 for BNT162b2) (Figure 1A and Supplementary Table S1). Interestingly, when compared with healthy subjects younger than 60 years of age who received BNT162b2 IM booster vaccination from our previous study [10] (GMC 2364, 95% CI 2006-2786 BAU/mL), both mRNA-1273 and BNT162b2 IM vaccination as well as mRNA-1273 ID vaccination generated similar or higher anti-RBD IgG concentrations, but lower concentrations by BNT162b2 ID vaccination.

**Figure 1.**
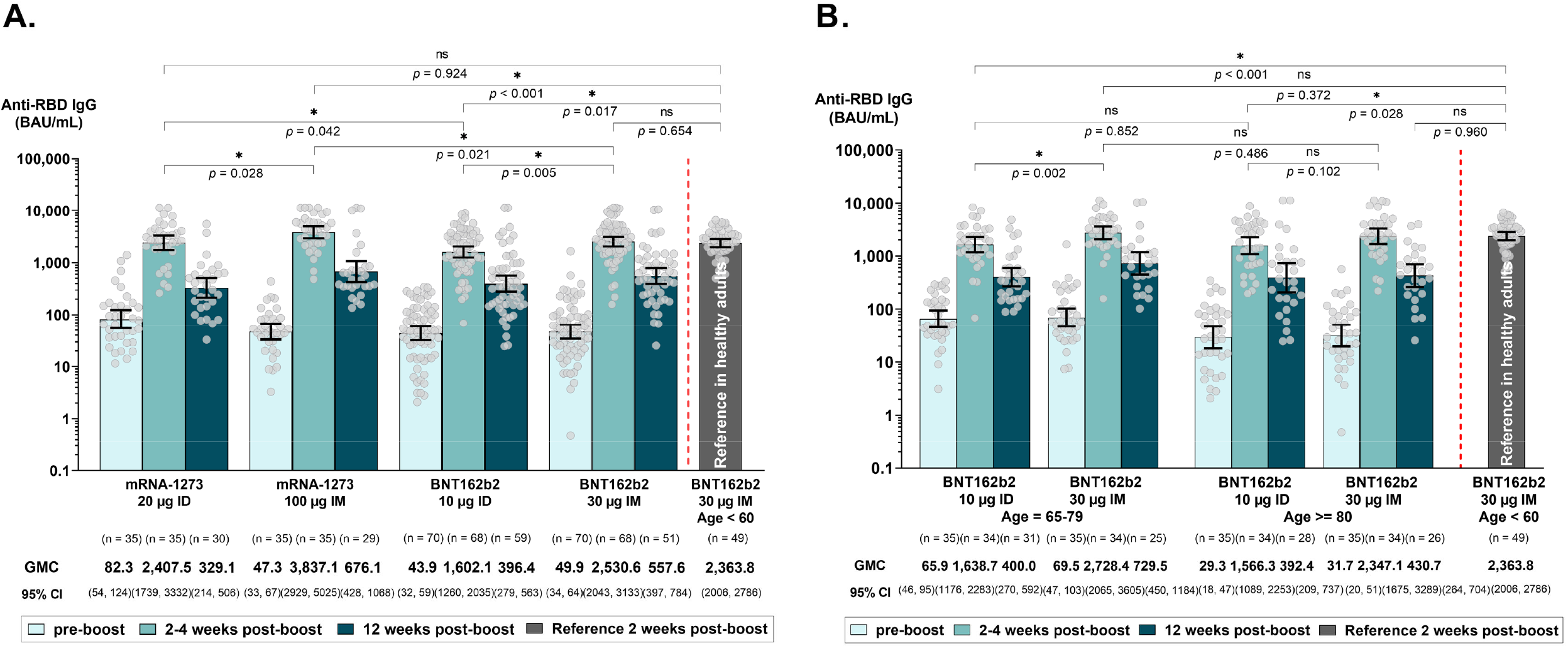
Anti-RBD IgG at baseline and following intramuscular or intradermal booster vaccination using mRNA-1273 or BNT162b2 vaccine **(A)** and in the BNT162b2 vaccine groups stratified by age (**B)**. Data presented as geometric mean concentration *(GMC)* with 95% confidence interval and compared using unpaired student t-test. Grey color bar presents the reference *GMC at 2 weeks after booster* in previous study among healthy adults age *<*60 years who received intramuscular BNT162b2 booster following 2-dose ChAdOx1 primary series [10].

Among participants in BNT162b2 groups, there were no significant differences in anti-RBD IgG responses between those who aged 65-79 years and those ≥80 years who received the similar route. When compared with reference group of younger adults aged <60 years, similar anti-RBD IgG responses were found for IM vaccination regardless of a those who aged 65-79 years or ≥80 years, whereas significantly lower responses were found for ID vaccination (Figure 1B and Supplementary Table S2).

At 12 weeks following the booster vaccination, the overall anti-RBD IgG GMC declined by 4-to 7-fold of the levels at 2-4 weeks; but were still between 4-to 14-fold higher than at baseline. The rate of decline as measured by GMRs between 12 weeks and 2-4 weeks post-boost were similar between the ID and IM groups, as well as in the different age range in the BNT162b2 groups.

### Neutralizing antibody responses against the SARS-CoV-2 variants

At baseline, only 95/210 (45.2%) and 11/210 (5.2%) participants had neutralizing antibodies (PVNT_50_) against Wuhan and Omicron BA.1. There was a significant increase in PVNT_50_ against ancestral Wuhan and Omicron variants following ID or IM booster vaccination; all participants became seropositive for Wuhan and around 89%, 95%, 88% of the study cohort were seropositive for Omicron BA.1, BA.2, and BA.4/5, respectively. Unlike what was seen for anti-RBD IgG, there was no statistical differences in PVNT_50_ against Wuhan strain between ID or IM of either vaccine [for mRNA-1273: ID 1,212 (95% CI 835,1759) vs IM 1,718 (95% CI 1130, 2612), *p*=0.210; for BNT162b2: ID 587 (95% CI 432, 798) vs IM: 712 (95% CI 502, 1009) *p*=0.407)]. However, both mRNA-1273 and BNT162b2 ID groups induced significantly lower PVNT_50_ for Omicron variants than IM of the same vaccine. In general, both mRNA-1273 ID or IM vaccination generated higher PVNT_50_ against Wuhan than ID or IM vaccination of BNT162b2 (*p*<0.01 for the comparisons of the same route), and ID mRNA1273 had similar GMT to IM BNT162b2 (*p*=0.577). When compared with the reference range for PVNT_50_ against Omicron BA.1 in younger adults (aged <60 years) who received BNT162b2 IM booster from our previous study, there was no significant differences for older people who received mRNA-1273 either by ID or IM, while significantly lower PVNT_50_ was observed for BNT162b2 groups (ID: GMT 147.8 or IM: 230.1 vs 521.2, *p*<0.001 and *p*=0.005, respectively) (Figure 2A and 2B and Supplementary Table S3).

**Figure 2.**
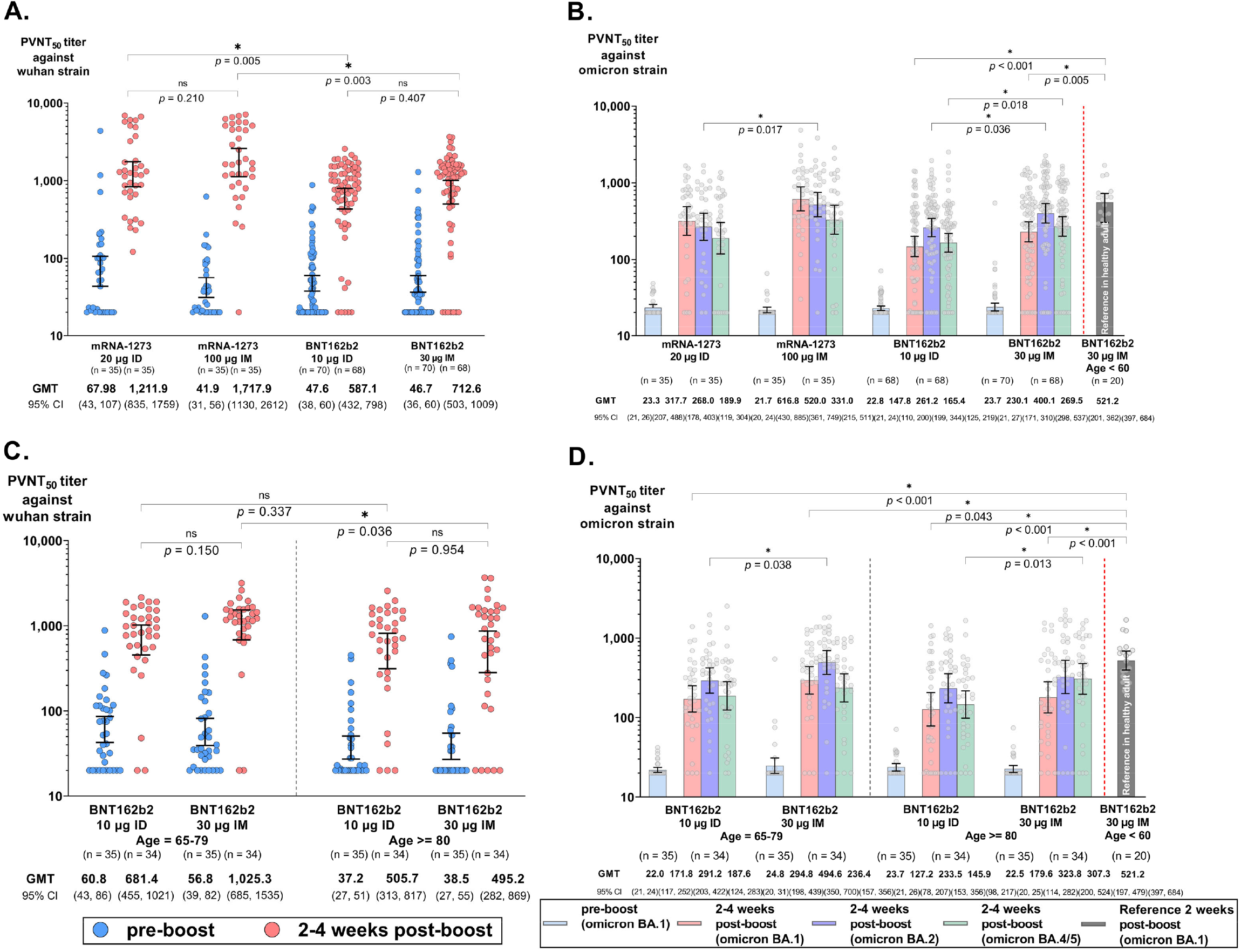
Neutralizing antibody titers (PVNT_*50*_) against SARS-CoV-2 ancestral Wuhan (A) and Omicron variants (B) at baseline and following intramuscular or intradermal booster vaccination using mRNA-1273 or BNT162b2 vaccine and in the BNT162b2 vaccine groups stratified by age **(C-D)**. Data presented as geometric mean neutralizing antibody titers (GMT) with 95% confidence interval (95%CI) and compared using unpaired student t-test. Grey color bar presents the reference PVNT_*50*_ GMT against Omicron BA.1 at 2 weeks after booster in previous study among healthy adults age <60 years who received intramuscular BNT162b2 booster following 2-dose ChAdOx1 primary series. The *p*-value displayed only if <0.05.

When stratified by participants aged 65-79 years and ≥80 years among the BNT162b2 groups following either route, the younger group generally had higher PVNT_50_ and proportion of seropositive against all strains, although this was only statistically significant for the PVNT_50_ against Wuhan following the IM route (GMT 1025.3 vs 495.2, *p*=0.036) (Figure 2C and 2D, and Supplementary Table S4).

### Cellular immune responses against ancestral Wuhan strain

Following the booster dose, all ID or IM vaccination groups generated significant increase in IFNγ-producing cell responses against the spike protein from baseline (Figure 3A and Supplementary Table S5). Participants who received ID vaccination had higher IFNγ responses than the respective IM route but was statistically significant only in mRNA-1273 vaccine (*p*=0.026). Interestingly, ID mRNA-1273 group had a significantly higher spike-specific IFNγ responses than IDBNT162b2 groups (*p*=0.047). There were no statistical differences in IFNγ response against NMO proteins between baseline and post booster for both mRNA-1273 and BNT162b2, as well as between ID and IM of either vaccine (Figure 3B and Supplementary Table S5).

**Figure 3.**
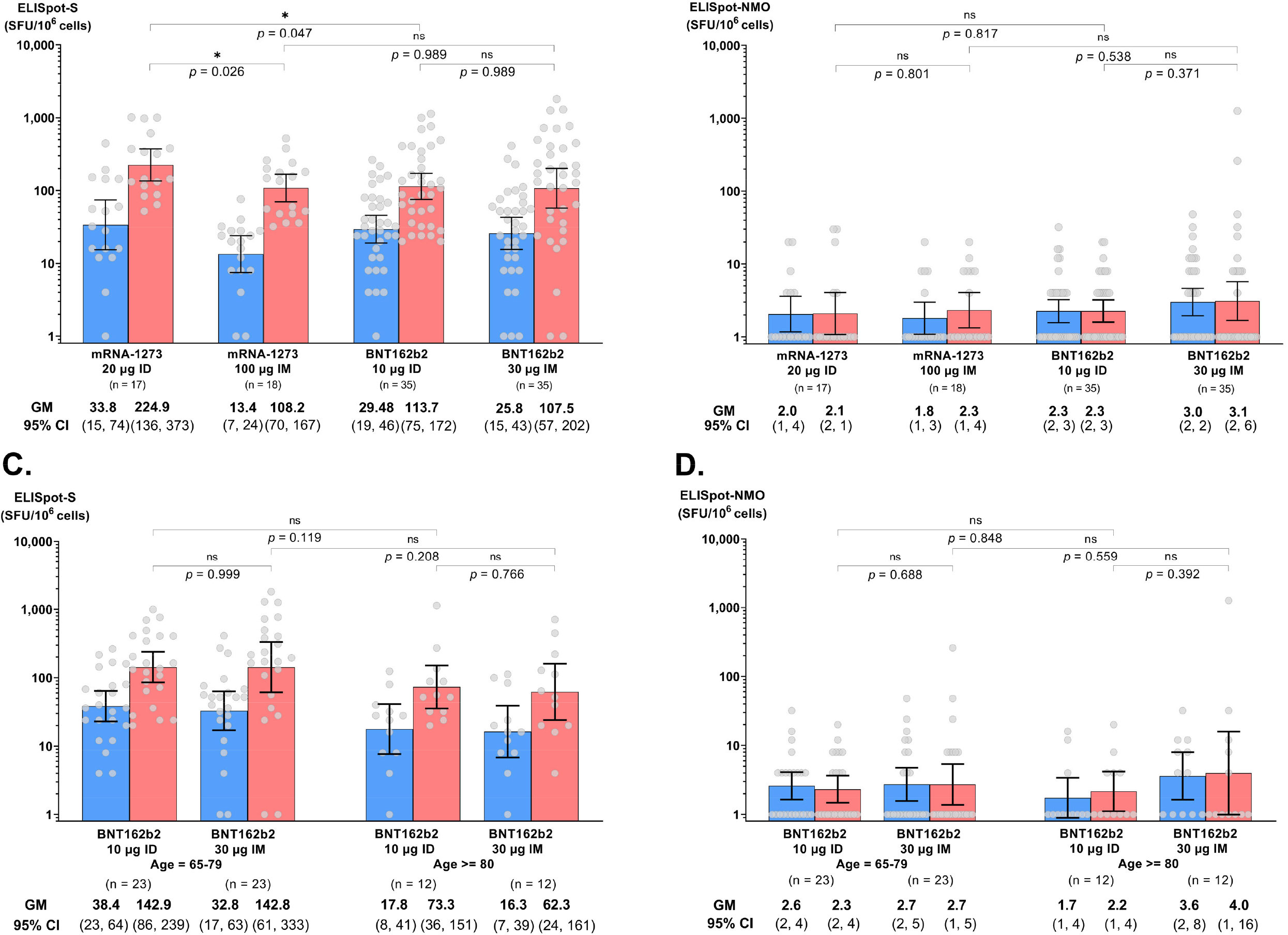
SARS-CoV-2 spike-specific IFNγ-producing cell responses (A) and NMO-specific IFNγ producing cell responses (B) at baseline and following intramuscular or intradermal booster vaccination using mRNA-1273 or BNT162b2 vaccine and in the BNT162b2 vaccine groups stratified by age **(C-D)**. Data presented as geometric mean of spot-forming unit/10^6^ cells and compared using student unpaired t-test.

When stratified by participants aged 65-79 years and ≥80 years among the BNT162b2 groups, similar IFNγ responses against spike protein was found between ID and IM vaccination within each age group. Between participants aged 65-79 years and participants aged ≥80 years, higher but not statistically significant IFNγ responses against the spike protein was observed. (Figure 3C and Supplementary Table S6). There were no statistical differences in IFNγ response against NMO proteins between ID and IM vaccination within each age group, and vaccination route between each age group (Figure 3D and Supplementary Table S6).

### Adverse events (AEs)

All AEs reported were mild (grade 1) to moderate (grade 2) and all participants recovered within 2-3 days. No serious AEs were found in this study (Figure 4 and Supplementary Table S7,S8).

**Figure 4.**
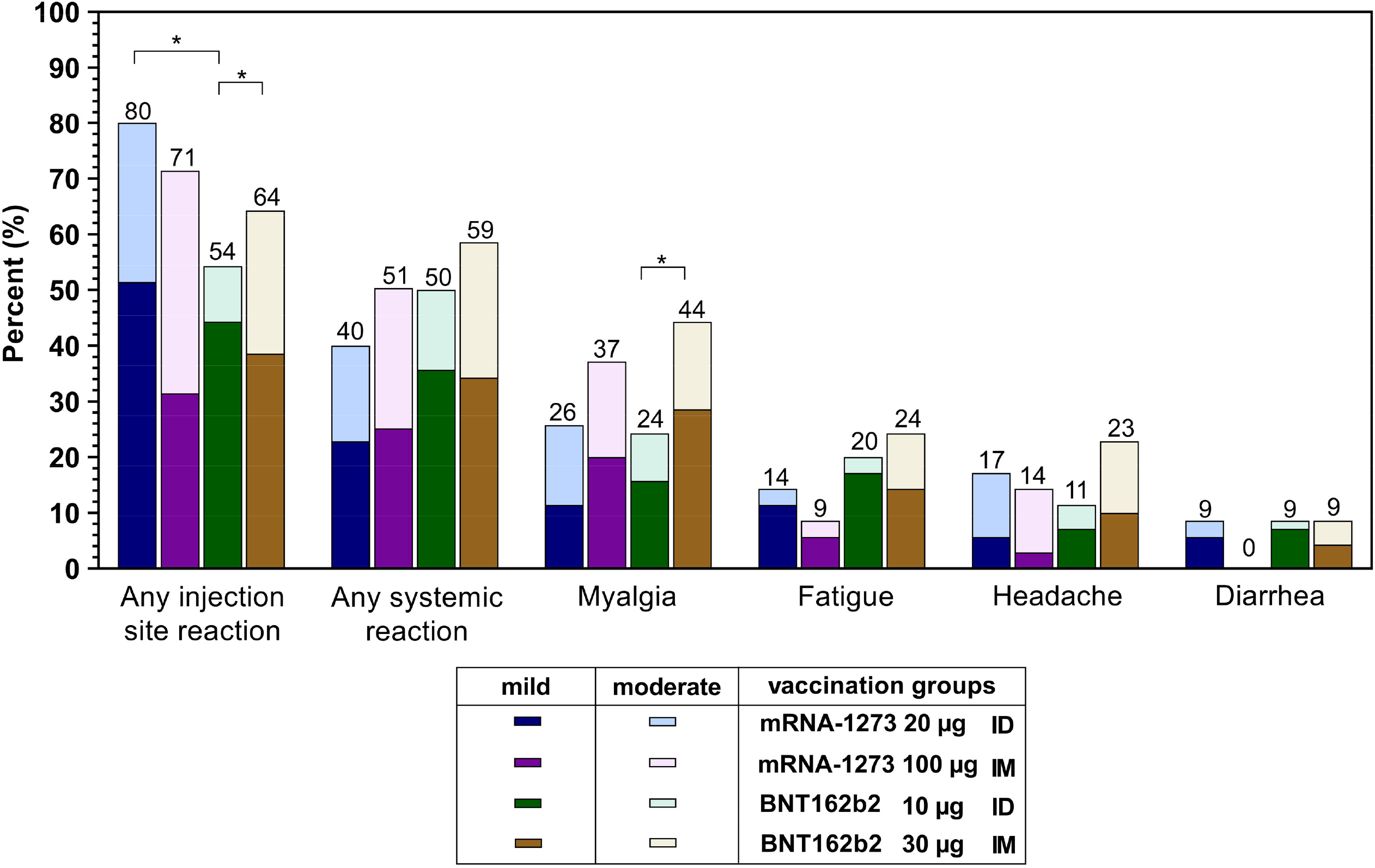
Adverse events reported in the 7 days following vaccination in the four study groups of intramuscular or intradermal route using mRNA-1273 or BNT162b2 vaccine. * = *p* <0.05.

The systemic AEs were not statistically different between the groups although IM groups tended to have a higher rate of AEs than ID (51% vs 40% for mRNA-1273, and 59% vs 50% for BNT162b2). Myalgia was more common with IM vaccination for both mRNA-1273 and BNT162b2 compared with ID vaccination. In contrast, ID route tend to have more frequent local AEs than IM for mRNA-1273 (80% vs 71%), whereas this was the opposite for BNT162b2 (54% vs 64%). mRNA-1273 in general induced more local reaction than BNT162b2 (*p* = 0.009).

## Discussion

To our knowledge, this is the first study that compare the immunogenicity and reactogenicity of fractional dose ID administration of mRNA vaccines as the booster vaccination in older adults with the standard-dose mRNA vaccines given IM route. We found that fractional dose (0.1 ml) of mRNA-1273 or BNT162b2 induced robust immunes responses, with mRNA-1273 mounting higher responses than BNT162b2. While lower antibody responses were observed between ID and IM vaccination of either mRNA-1273 and BNT162b2, fractional dose mRNA-1273 ID vaccination generally induced similar or higher antibody and cellular responses as those induced by standard dose BNT162b2 IM vaccination. In addition, the antibody responses induced by ID and IM vaccinations appear to wane at a similar rate. Local adverse events were reportedly more common following ID vaccination than IM vaccination, but the systemic adverse events were less frequent following ID vaccination. Overall, fractional ID vaccination of mRNA vaccine may be considered as an alternative to standard IM vaccination for older adults, particularly in settings where there are vaccine shortages (e.g. new COVID-19 vaccines) and/or vaccine hesitancy due to high reactogenicity.

Older adults are the primary target for COVID-19 vaccination as they have increased risk of severe COVID-19 due to immunosenecence and likely have comorbidities. Previous studies of COVID-19 booster vaccination in older adults given via IM route were found to be highly effective against COVID-19, with more than 80% against any COVID-19-related symptoms and more than 90% against hospitalization and death, prior to the emergence of SARS-CoV-2 omicron variants [11-13]. Neutralizing antibodies are thought to be the primary mechanism of protection against SARS-CoV-2 infection [14], while cellular immune responses are thought to be more important against severe COVID-19 [15]. In our study, despite the lower antibody response and similar or higher cellular immune responses found following ID administration when compared with respective IM administration of the same vaccine, the clinical relevance is unknown. It is possible that ID administration may offer similar protection as with IM administration. The higher immunogenicity following mRNA-1273 IM vaccination than BNT162b2 IM vaccination is consistent with previous studies. A higher antigen amount in mRNA-1273 than in BNT162b2 is likely to account for this finding. However, it is important to note that the amount of vaccine content used in the ID administration of fractional mRNA-1273 dose is less than half of the current recommended IM administration dose (50µg). Whether the immune responses induced by fractional dose given via ID route induced a similar immune memory cell responses as standard dose IM delivery remains to be determined.

We observed marginally lower neutralizing antibodies and cellular immune responses among adults aged ≥80 years compared with adults aged 65-79 years. This is consistent with previous studies of BNT162b2 that found lower neutralizing antibody responses in older adults compared with younger adults [16,17]. This underscores the importance of choice of vaccine in older adults. Nevertheless, our findings suggest that older adults can mount robust immunity following a booster dose, even with ID administration of fractional dosage.

Consistent with our findings from previous studies on ID administration, we found lower systemic adverse effects but higher local adverse effects with ID route when compared with IM delivery. This could lead to lower vaccine hesitancy in older people. Of note, we found the proportions of AEs reported following ID route in this study were lower than that report in adults younger <60 years (54-80% vs 91% for local AEs, and 40-50% vs 69%) [7]. This is consistent with other studies that found lower reactogenicity in older people [18].

There are some limitations in this study. Firstly, the sample size is small and made up of largely females, which may have influenced our ability to detect potential differences between study groups. Secondly, this was an open label study since the administration route cannot be concealed, which may introduce bias on reporting of adverse events, although this is unlikely to affect our immunogenicity findings. Thirdly, data from younger adults were based on another study cohort which may not be directly comparable. However, the study was conducted under similar settings and the data were generated using the same testing methods by the same laboratory, minimizing variability. Lastly, our data may not be generalizable to other COVID-19 vaccines and other populations such as those with other comorbidities

In conclusion, we demonstrated that fractional ID mRNA-1273 or BNT162b2 booster vaccination generate robust immune responses and lower systemic AEs in older adults, ID despite inducing lower antibody responses when compared with standard dose IM vaccination. ID route could be considered an alternative option for older people. Further investigation of vaccine efficacy of ID vaccination is warranted.

## Data Availability

All data produced in the present study are available upon reasonable request to the authors.

## Acknowledgements

The authors wish to thank all of the participants for their collaboration. The authors also would like to express the gratitude to Siriraj Geriatric Clinic staff (Dujpratana Pisalsarakij, Napaporn Pengsorn, Pensri Chaopanitwet, Monthira Thammasalee, Pitiporn Siritipakorn, and Wiyachatr Monklang), SICRES staff (Chatkamol Pheerapanyawaranun, Laddawan Jansarikit, Thiranuch Wongsawat, and Suparat Atakulreka), and Siriraj Department of Immunology staff (Utane Rungpanich, Pinklow Umrod, Jintapa Sueasuay, Therapit Butlop, Winita Viriyakijja, Maneeprang Towarapa, Kotchamon Chuaykaew, Tanyawan Saenwad, Jirapond Boonma, Chayaporn Janthanong, Naharuthai Inthasin, Rawipas Saisuwan, and Nuttawan Kassaket) who give great contribution to the study.

