## Supplementary Table S1 for "Immunogenicity and reactogenicity of mRNA COVID-19 vaccine booster administered by intradermal or intramuscular route in Thai Older adults"

**Supplementary Table S1.** Anti-SARS-CoV-2 receptor binding domain (anti-RBD) IgG antibody concentrations among older people aged ≥ 65 years at 12-24 weeks after 2-dose of ChAdOx1 vaccination (pre-boost) and at 2-4 weeks after the booster (3^rd^ dose) vaccination (2-4 weeks and 12 weeks post-boost) by type and route of booster vaccine administration

| **Level of anti-RBD IgG (BAU/mL)** | **Type and route of booster vaccine administration** | | | | | ***p*-value** |
| --- | --- | --- | --- | --- | --- | --- |
|  | **Total** | **mRNA-1273 (ID)** | **mRNA-1273 (IM)** | **BNT162b2 (ID)** | **BNT162b2 (IM)** |  |
| **Pre-boost (12-24 weeks after primary series)** | | | | | | |
| **Number of participants** | **(n = 210)** | **(n = 35)** | **(n = 35)** | **(n = 70)** | **(n = 70)** | ***p*-value** |
| **GMC at pre-boost (95% CI)** | 50.50  (42.61,  59.84) | 82.34  (54.78, 123.78) | 47.30  (33.30,  67.19) | 43.92  (32.31, 59.70) | 49.97  (34.41, 64.13) | 0.106 |
| **2-4 weeks post-boost** | | | | | | |
| **Number of participants** | **(n = 206)** | **(n = 35)** | **(n = 35)** | **(n = 68)** | **(n = 68)** | ***p*-value** |
| **GMC at 2-4 weeks**  **post-boost (95% CI)** | 2,315.92  (2030.41, 2641.57) | 2,407.53  (1739.47, 3332.16) | 3,837.12  (2929.87, 5025.30) | 1,602.10  (1260.78, 2035.82) | 2,530.58  (2043.51, 3133.75) | <0.001* |
| Aggregate GMR: ID and IM  at 2-4 weeks post-boost (95% CI) | | 0.82  (0.68, 0.98) | | 0.82  (0.71, 0.94) | |  |
| *p*-value | | 0.028* | | 0.005* | |  |
| **GMR between 2-4 weeks post-boost and pre-boost**  **(95% CI)** | **45.11**  **(38.82,**  **52.41)** | **29.24**  **(20.76, 41.19)** | **81.12**  **(60.92, 108.03)** | **35.31**  **(27.53, 45.28)** | **53.26**  **(40.12, 70.69)** | **<0.001*** |
| **12 weeks post-boost** | | | | | | |
| **Number of participants** | **(n = 169)** | **(n = 30)** | **(n = 29)** | **(n = 59)** | **(n = 51)** | ***p*-value** |
| **GMC at 12 weeks**  **post-boost (95% CI)** | 465.92  (384.49,  564.58) | 329.10  (213.95,  506.22) | 676.11  (427.89,  1068.32) | 396.40  (278.99,  563.21) | 557.63  (396.51,  784.24) |  |
| Aggregate GMR: ID and IM  at 12 weeks post-boost (95% CI) | | 0.73  (0.56, 0.96) | | 0.86  (0.70, 1.07) | |  |
| *p*-value | | 0.022* | | 0.168 | |  |
| **GMR between 12 weeks post-boost and 2-4 weeks post-boost (95% CI)** | **0.21**  **(0.17,**  **0.24)** | **0.13**  **(0.10,**  **0.18)** | **0.18**  **(0.11,**  **0.29)** | **0.25**  **(0.19,**  **0.34)** | **0.23**  **(0.17,**  **0.29)** | **0.048** |

Note: * *p* ≤ 0.05

Abbreviation: GMC: geometric mean concentration, BAU: binding antibody unit, GMR: geometric mean ratio
