## Supplementary Table S2 for "Immunogenicity and reactogenicity of mRNA COVID-19 vaccine booster administered by intradermal or intramuscular route in Thai Older adults"

**Supplementary Table S2.** Anti-SARS-CoV-2 receptor binding domain (anti-RBD) IgG antibody concentrations among older people at 12-24 weeks after 2-dose of ChAdOx1 vaccination (pre-boost) and at 2-4 weeks after the booster (3^rd^ dose) vaccination (2-4 weeks and 12 weeks post-boost) by routes of BNT162b2 vaccination, and by age range

| **Level of anti-RBD IgG (BAU/mL)** | **Type and route of booster** **vaccine administration** | | | | | **p-value** |
| --- | --- | --- | --- | --- | --- | --- |
|  | **Total** | **BNT162b2**  **(ID)**  **Age 65-79** | **BNT162b2**  **(ID)**  **Age ≥ 80** | **BNT162b2**  **(IM)**  **Age 65-79** | **BNT162b2**  **(IM)**  **Age ≥ 80** |  |
| **Pre-boost (12-24 weeks after primary series)** | | | | | | |
| **Number of participants** | **(n = 140)** | **(n = 35)** | **(n = 35)** | **(n = 35)** | **(n = 35)** | ***p* -value** |
| **GMC at pre-boost**  **(95% CI)** | 45.42  (36.60, 56.37) | 65.89  (45.73, 94.94) | 29.28  (18.25, 46.97) | 69.53  (47.15, 102.54) | 31.73  (19.85, 50.72) | 0.189 |
| **2-4 weeks post-boost** | | | | | | |
| **Number of participants** | **(n = 136)** | **(n = 34)** | **(n = 34)** | **(n = 34)** | **(n = 34)** | ***p*-value** |
| **GMC at 2-4 weeks post-boost**  **(95% CI)** | 2,013.51  (1710.34,  2370.43) | 1,638.69  (1176.07, 2283.27) | 1,566.32  (1089.17, 2252.51) | 2,728.42  (2065.01, 3604.97) | 2,347.08  (1675.09, 3288.65) | 0.042* |
| Aggregate GMR: age 65-79 and age ≥ 80 at 2-4 weeks post-boost (95% CI) | | 1.02  (0.89, 1.29) | | 1.07  (0.89, 1.29) | |  |
| *p*-value | | 0.852 | | 0.486 | |  |
| **GMR between 2-4 weeks post boost and pre-boost (95% CI)** | 30.59  (24.58, 38.08) | 23.79  (17.58, 32.19) | 52.40  (36.53, 75.19) | 39.36  (28.93, 53.54) | 72.06  (45.05, 115.28) | 0.021* |
| **12 weeks post-boost** | | | | | | |
| **Number of participants** | **(n = 110)** | **(n = 31)** | **(n = 28)** | **(n = 25)** | **(n = 26)** | **p-value** |
| **GMC at 12 weeks post-boost (95% CI)** | 464.36  (363.74,  592.81) | 400.01  (270.41, 591.71) | 392.44  (209.10, 736.52) | 729.51  (449.67,  1183.51) | 430.68  (263.51,  703.89) | 0.260 |
| Aggregate GMR: age 65-79 and age ≥ 80 at 2-4 weeks post-boost (95% CI) | | 1.01  (0.74, 1.37) | | 1.26  (0.94, 1.68) | |  |
| p-value | | 0.957 | | 0.122 | |  |
| **GMR between 12 weeks post boost and 2-4 weeks post-boost (95% CI)** | 0.24  (0.20,  0.29) | 0.26  (0.17,  0.39) | 0.25  (0.15,  0.40) | 0.26  (0.18,  0.38) | 0.20  (0.13,  0.29) | 0.772 |

Note: * *p* < 0.05

Abbreviation: IM: Intramuscular administration, ID: intradermal administration, GMC: geometric mean concentration, BAU/mL: binding antibody unit per milliliter, GMR: geometric mean ratio, CI: confidence interval
