## Supplementary Table S3 for "Immunogenicity and reactogenicity of mRNA COVID-19 vaccine booster administered by intradermal or intramuscular route in Thai Older adults"

**Supplementary Table S3.** Pseudovirus Neutralization Titer (PVNT_50_) titres among older people aged ≥65 years at 12-24 weeks after 2-dose of ChAdOx1 vaccination (pre-boost) and at 2-4 weeks after the booster (3^rd^ dose) vaccination (2-4 weeks post-boost) by type and route of booster vaccine administration

| **Pseudovirus Neutralization Assay (PVNT_50_)** | **Type and route of booster** **vaccine administration** | | | | | | ***p*-value** |
| --- | --- | --- | --- | --- | --- | --- | --- |
|  | **Total** | **mRNA-1273**  **(ID)** | | **mRNA-1273**  **(IM)** | **BNT162b2 (ID)** | **BNT162b2 (IM)** |  |
| **Pre-boost (12-24 weeks after primary series)** | | | | | | | |
| **Number of participants** | **(n = 210)** | **(n = 35)** | | **(n = 35)** | **(n = 70)** | **(n = 70)** | ***p*-value** |
| **GMT against Wuhan strain at pre-boost (95%CI)** | 49.13  (42.54, 56.73) | 67.98  (43.35,  106.63) | | 41.86  (31.12,  56.30) | 47.56  (37.54,  60.24) | 46.73  (36.31,  60.16) | 0.229 |
| **GMT against Omicron BA.1 strain at pre-boost (95%CI)** | 22.98  (21.88, 24.13) | 23.29  (21.15,  25.64) | | 21.65  (19.89,  23.56) | 22.83  (21.43,  24.33) | 23.66  (20.99,  26.66) | 0.688 |
| PVNT_50_ ≥ 1:40 against Omicron BA.1 at pre-boost, n (%) | 11  (5.24) | 2  (5.71) | | 1  (2.86) | 2  (2.86) | 6  (8.57) | 0.463 |
| **2-4 weeks post-boost** | | | | | | |  |
| **Number of participants** | **(n = 206)** | **(n = 35)** | | **(n = 35)** | **(n = 68)** | **(n = 68)** | ***p*-value** |
| **GMT against Wuhan strain at 2-4 weeks post-booster (95%CI)** | 849.53  (706.29,  1021.82) | 1,211.98  (835.08, 1759.00) | | 1,717.94  (1129.87,  2612.11) | 587.07  (431.74,  798.30) | 712.56  (503.01,  1009.40) | <0.001* |
| Aggregate GMR: ID and IM against Wuhan strain at 2-4 weeks post-boost (95%CI) | | 0.86  (0.68, 1.09) | | | 0.92  (0.75, 1.12) | |  |
| *p*-value | | 0.210 | | | 0.407 | |  |
| **GMT against Omicron BA.1 strain at 2-4 weeks post-boost (95%CI)** | 248.32  (207.62,  297.00) | 317.67  (206.69,  488.24) | | 616.75  (429.86,  884.88) | 147.81  (109.71,  200.13) | 230.10  (170.56,  310.43) | <0.001* |
| PVNT_50_ ≥ 1:40 against Omicron BA.1 at 2-4 weeks post-boost, n (%) | 184  (89.32) | 31  (88.57) | | 34  (97.14) | 57  (83.82) | 62  (91.18) | 0.211 |
| Aggregate GMR: ID and IM against Omicron BA.1 strain at 2-4 weeks  post-boost (95%CI) | | 0.75  (0.59, 0.95) | | | 0.83  (0.69, 0.98) | |  |
| *p*-value | | 0.019* | | | 0.040* | |  |
| **GMT against Omicron BA.2 strain at 2-4 weeks post-boost (95%CI)** | 339.49  (288.70,  399.22) | 268.02  (178.16,  403.18) | | 519.99  (360.87,  749.29) | 261.20  (198.64,  343.45) | 400.16  (298.47,  536.50) | 0.013* |
| PVNT_50_ ≥ 1:40 against Omicron BA.2 at 2-4 weeks post-boost, n (%) | 195  (95.12) | 32  (94.12) | | 34  (97.14) | 64  (94.12) | 65  (95.59) | 0.914 |
| Aggregate GMR: ID and IM against Omicron BA.2 strain at 2-4 weeks  post-boost (95%CI) | | 0.75  (0.59, 0.95) | | | 0.83  (0.70, 0.99) | |  |
| *p*-value | | 0.017* | | | 0.036* | |  |
| **GMT against Omicron BA.4/5 strain at 2-4 weeks post-boost (95%CI)** | 223.81  (188.42,  265.85) | 189.88  (118.69,  303.78) | | 331.01  (214.51,  510.78) | 165.38  (125.03,  218.75) | 269.49  (200.58,  362.07) | 0.023* |
| PVNT_50_ ≥ 1:40 against Omicron BA.4/5 at 2-4 weeks post-boost,  n (%) | 181  (87.86) | 28  (80.00) | | 31  (88.57) | 60  (88.24) | 62  (91.18) | 0.446 |
| Aggregate GMR: ID and IM against Omicron BA.4/5 strain at 2-4 weeks post-boost (95%CI) | | 0.79  (0.60, 1.03) | | | 0.81  (0.68, 0.96) | |  |
| *p*-value | | 0.082 | | | 0.018* | |  |
| **GMR: Wuhan strain between 2-4 weeks post-boost and**  **pre-boost (95%CI)** | **17.07**  **(13.90,**  **20.96)** | **17.83**  **(12.02,**  **26.44)** | **41.04**  **(27.94,**  **60.30)** | | **12.03**  **(8.05,**  **17.98)** | **15.07**  **(10.49,**  **21.66)** | **<0.001*** |
| **GMR: Omicron BA.1 strain between 2-4 weeks post-boost  and pre-boost (95%CI)** | **10.78**  **(8.98,**  **12.93)** | **13.64**  **(8.59,**  **21.66)** | **28.49**  **(19.89,**  **40.81)** | | **6.45**  **(4.75,**  **8.76)** | **9.68**  **(7.20,**  **13.02)** | **<0.001*** |

Note: * *p* ≤ 0.05

Abbreviation: GMC: geometric mean concentration, BAU: binding antibody unit, GMR: geometric mean ratio

Titer values reported as below the lower limit of detection (LLOD = 1:40) were replaced with 20
