## Supplementary Table S4 for "Immunogenicity and reactogenicity of mRNA COVID-19 vaccine booster administered by intradermal or intramuscular route in Thai Older adults"

**Supplementary Table S4.** Pseudovirus Neutralization Titer (PVNT_50_) among older people at 12-24 weeks after 2-dose of ChAdOx1 vaccination (pre-boost) and at 2-4 weeks after the booster (3^rd^ dose) vaccination (2-4 weeks post-boost) by type and by routes of BNT162b2 vaccination, and by age range

| **Pseudovirus Neutralization Assay (PVNT_50_)** | **Type and route of booster** **vaccine administration** | | | | | ***p*-value** |
| --- | --- | --- | --- | --- | --- | --- |
|  | **Total** | **BNT162b2**  **(ID)**  **Age 65-79** | **BNT162b2**  **(ID)**  **Age ≥ 80** | **BNT162b2**  **(IM)**  **Age 65-79** | **BNT162b2**  **(IM)**  **Age ≥ 80** |  |
| **Pre-boost (12-24 weeks after primary series)** | | | | | | |
| **Number of participants** | **(n = 140)** | **(n = 35)** | **(n = 35)** | **(n = 35)** | **(n = 35)** | **p-value** |
| **GMT against Wuhan strain**  **at pre-boost (95%CI)** | 47.14  (39.74,  55.93) | 60.80  (42.80,  86.37) | 37.20  (27.21,  50.84) | 56.80  (39.46,  81.76) | 38.45  (26.94,  54.88) | 0.084 |
| **GMT against Omicron BA.1 strain**  **at pre-boost (95%CI)** | 23.24  (21.74,  24.85) | 21.98  (20.47,  23.60) | 23.73  (21.32,  26.41) | 24.82  (19.87,  30.99) | 22.55  (20.39,  24.95) | 0.595 |
| PVNT_50_ _≥_ 1:40 against omicron BA.1  at pre-boost, n (%) | 8  (5.71) | 1  (2.86) | 1  (2.86) | 4  (11.43) | 2  (5.71) | 0.527 |
| **2-4 weeks post-boost** | | | | | | |
| **Number of participants** | **(n = 136)** | **(n = 34)** | **(n = 34)** | **(n = 34)** | **(n = 34)** | **p-value** |
| **GMT against Wuhan strain**  **at 2-4 weeks post-boost (95%CI)** | 646.78  (513.97,  813.91) | 681.49  (454.96,  1020.81) | 505.74  (313.19,  816.66) | 1,025.32  (685.09,  1534.51) | 495.20  (282.30,  868.68) | 0.091 |
| Aggregate GMR: ID and IM against Wuhan strain  at 2-4 weeks post-boost (95%CI) | | 1.14  (0.87, 1.49) | | 1.37  (1.02, 1.82) | |  |
| *p*-value | | 0.337 | | 0.036* | |  |
| **GMT against Omicron BA.1 strain**  **at 2-4 weeks post-boost (95%CI)** | 184.42  (148.95,  228.34) | 171.77  (117.30,  251.53) | 127.20  (78.24,  206.81) | 294.83  (198.13,  438.72) | 179.59  (114.20,  282.40) | 0.049* |
| PVNT_50_ ≥ 1:40 against omicron BA.1 strain at 2-4 weeks post-boost, n (%) | 119  (87.50) | 31  (91.18) | 26  (76.47) | 32  (94.12) | 30  (88.24) | 0.048* |
| Aggregate GMR: ID and IM against Omicron BA.1 strain at 2-4 weeks post-boost (95%CI) | | 1.14  (0.88, 1.48) | | 1.24  (0.96, 1.60) | |  |
| *p*-value | | 0.326 | | 0.099 | |  |
| **GMT against Omicron BA.2 strain**  **at 2-4 weeks post-boost (95%CI)** | 323.30  (264.35,  395.39) | 291.24  (202.54,  421.65) | 233.46  (152.99,  356.25) | 494.60  (349.67,  699.60) | 323.76  (200.18,  523.63) | 0.065 |
| PVNT_50_ ≥ 1:40 against omicron BA.2 strain at 2-4 weeks post-boost, n (%) | 129  (94.85) | 33  (97.06) | 31  (91.18) | 34  (100.00) | 31  (91.18) | 0.766 |
| Aggregate GMR: ID and IM against Omicron  BA.2 strain at 2-4 weeks post-boost (95%CI) | | 1.10  (0.87, 1.40) | | 1.20  (0.93, 1.55) | |  |
| *p*-value | | 0.417 | | 0.151 | |  |
| **GMT against Omicron BA.4/5 strain**  **at 2-4 weeks post-boost (95%CI)** | 210.55  (158.57,  279.57) | 187.55  (124.34,  282.89) | 145.83  (98.10,  216.78) | 236.37  (157.10,  355.64) | 307.25  (197.18,  478.76) | 0.068 |
| PVNT_50_ ≥ 1:40 against omicron BA.4/5 strain at 2-4 weeks post-boost, n (%) | 122  (89.71) | 29  (85.29) | 31  (91.18) | 31  (91.18) | 31  (91.18) | 0.891 |
| Aggregate GMR: ID and IM against Omicron BA.4/5 strain at 2-4 weeks post-boost (95%CI) |  | 1.12  (0.87, 1.42) | | 0.89  (0.69, 1.15) | |  |
| *p*-value |  | 0.373 | | 0.380 | |  |
| **GMR: Wuhan strain between 2-4 weeks post-boost and pre-boost (95%CI)** | **13.97**  **(9.59,**  **20.35)** | **10.85**  **(6.01,**  **19.58)** | **13.35**  **(7.52,**  **23.69)** | **17.98**  **(11.10,**  **29.13)** | **12.63**  **(7.21,**  **22.13)** | **0.612** |
| **GMR: Omicron BA.1 strain between 2-4 weeks post-boost and pre-boost (95%CI)** | **9.59**  **(7.38,**  **12.47)** | **7.79**  **(5.41,**  **11.22)** | **5.33**  **(3.23,**  **8.82)** | **11.81**  **(8.05,**  **17.32)** | **7.93**  **(4.99,**  **12.60)** | **0.077** |

Note: * *p* ≤ 0.05

Abbreviation: GMC: geometric mean concentration, BAU: binding antibody unit, GMR: geometric mean ratio

Titer values reported as below the lower limit of detection (LLOD = 1:40) were replaced with 20
