## Supplementary Table S5 for "Immunogenicity and reactogenicity of mRNA COVID-19 vaccine booster administered by intradermal or intramuscular route in Thai Older adults"

**Supplementary Table S5.** ELISpot-S and NMO among older people aged >65 years at 12-24 weeks after 2-dose of ChAdOx1 vaccination (pre-boost) and at 2-4 weeks after the booster (3^rd^ dose) vaccination (2-4 weeks post-boost) by type and route of booster vaccine administration.

| **ELISpot response**  **(SFU/10^6^ cells)** | **Type and route of booster** **vaccine administration** | | | | | ***p*-value** |
| --- | --- | --- | --- | --- | --- | --- |
|  | **Total** | **mRNA-1273**  **(ID)** | **mRNA-1273**  **(IM)** | **BNT162b2**  **(ID)** | **BNT162b2 (IM)** |  |
| **Pre-boost (12-24 weeks after primary series)** | | | | | | |
| **Number of participants** | **(n = 105)** | **(n = 17)** | **(n = 18)** | **(n = 35)** | **(n = 35)** | **p-value** |
| ELISpot-S  GM at pre-boost (95%CI) | 25.19  (19.25, 32.97) | 33.82  (15.36, 74.46) | 13.40  (7.48, 24.00) | 29.48  (19.05, 45.63) | 25.82  (15.48, 43.06) | 0.176 |
| ELISpot-NMO  GM at pre-boost  (95%CI) | 2.35  (1.89, 2.93) | 2.05  (1.17, 3.61) | 1.80  (1.09, 2.99) | 2.26  (1.57, 3.25) | 3.00  (1.95, 4.63) | 0.456 |
| **2-4 weeks post-boost** | | | | | |  |
| **Number of participants** | **(n = 105)** | **(n = 17)** | **(n = 18)** | **(n = 35)** | **(n = 35)** | ***p*-value** |
| ELISpot-S  GM at 2-4 weeks  post-boost (95%CI) | 123.54  (94.53,  161.46) | 224.89  (135.67,  372.77) | 108.15  (69.84,  167.48) | 113.69  (75.30,  171.66) | 107.46  (57.27,  201.64) | 0.282 |
| Aggregate GMR: ID and IM of ELISpot-S at 2-4 weeks post-boost (95%CI) | | 1.37  (1.04, 1.82) | | 1.02  (0.74, 1.41) | |  |
| *p*-value | | 0.026* | | 0.989 | |  |
| ELISpot-NMO  GM at 2-4 weeks  post-boost (95%CI) | 2.49  (1.91, 3.25) | 2.09  (1.08, 4.07) | 2.32  (1.33, 4.07) | 2.26  (1.59, 3.22) | 3.10  (1.68, 5.72) | 0.717 |
| Aggregate GMR: ID and IM of  ELISpot-NMO at 2-4 weeks post-boost (95%CI) | | 0.96  (0.67, 1.37) | | 0.87  (0.65, 1.18) | |  |
| *p*-value | | 0.801 | | 0.371 | |  |
| **ELISpot-S**  **GMR at 2-4 weeks post-boost and pre-boost (95%CI)** | **4.90**  **(3.75, 6.42)** | **6.65**  **(3.65, 12.10)** | **8.07**  **(3.96, 16.47)** | **3.86**  **(2.58, 5.76)** | **4.16**  **(2.39, 7.26)** | **0.199** |
| **ELISpot-NMO**  **GMR at 2-4 weeks post-boost and pre-boost (95%CI)** | **1.06**  **(0.81, 1.38)** | **1.02**  **(0.65, 1.60)** | **1.29**  **(0.83, 2.00)** | **1.00**  **(0.61, 1.63)** | **1.03**  **(0.57, 1.87)** | **0.932** |

Note: * *p* ≤ 0.05

Abbreviation: GM: geometric mean, SFU/10^6^ cells: spot forming units per million cells, GMR: geometric mean ratio
