## Supplementary Table S6 for "Immunogenicity and reactogenicity of mRNA COVID-19 vaccine booster administered by intradermal or intramuscular route in Thai Older adults"

**Supplementary Table S6.** ELISpot-S and NMO among older people at 12-24 weeks after 2-dose of ChAdOx1 vaccination (pre-boost) and at 2-4 weeks after the booster (3rd dose) vaccination (2-4 weeks post-boost) by type and routes of BNT162b2 vaccination, and by age range

| **ELISpot responses**  **(SFU/10^6^ cells)** | **Type and route of booster** **vaccine administration** | | | | | ***p*-value** |
| --- | --- | --- | --- | --- | --- | --- |
|  | **Total** | **BNT162b2**  **(ID)**  **Age 65-79** | **BNT162b2**  **(ID)**  **Age ≥ 80** | **BNT162b2**  **(IM)**  **Age 65-79** | **BNT162b2**  **(IM)**  **Age ≥ 80** |  |
| **Pre-boost (12-24 weeks after primary series)** | | | | | | |
| **Number of participants** | **(n = 70)** | **(n = 23)** | **(n = 12)** | **(n = 23)** | **(n = 12)** | ***p*-value** |
| **ELISpot-S**  **GM at pre-boost (95%CI)** | 27.59  (19.87,  38.30) | 38.42  (23.01,  64.14) | 17.75  (7.65,  41.21) | 32.81  (17.02,  63.28) | 16.31  (6.82,  39.05) | 0.198 |
| **ELISpot-NMO**  **GM at pre-boost (95%CI)** | 2.60  (1.97,  3.44) | 2.59  (1.64,  4.10) | 1.74  (0.89,  3.39) | 2.73  (1.57,  4.76) | 3.60  (1.63,  7.96) | 0.501 |
| **2-4 weeks post-boost** | | | | | | |
| **Number of participants** | **(n = 70)** | **(n = 23)** | **(n = 12)** | **(n = 23)** | **(n = 12)** | ***p*-value** |
| **ELISpot-S**  **GM at 2-4 weeks post-boost (95%CI)** | 110.53  (76.61,  159.48) | 142.93  (85.45,  239.07) | 73.32  (35.55,  151.21) | 142.83  (61.28,  332.91) | 62.29  (24.15,  160.62) | 0.287 |
| Aggregate GMR: ID and IM of ELISpot-S at 2-4 weeks post-boost (95%CI) | | 1.34  (0.92, 1.93) | | 1.43  (0.81, 2.54) | |  |
| *p*-value | | 0.120 | | 0.208 | |  |
| ELISpot-NMO  GM at 2-4 weeks post-boost (95%CI) | 2.65  (1.87,  3.74) | 2.32  (1.48,  3.64) | 2.16  (1.11,  4.19) | 2.72  (1.38,  5.37) | 3.97  (0.99, 15.92) | 0.721 |
| Aggregate GMR: ID and IM of ELISpot-NMO at 2-4 weeks post-boost (95%CI) | | 1.03  (0.74, 1.43) | | 0.85  (0.48, 1.50) | |  |
| *p*-value | | 0.848 | | 0.559 | |  |
| **ELISpot-S**  **GMR at 2-4 weeks post-boost and pre-boost (95%CI)** | **5.18**  **(3.79,**  **7.08)** | **3.72**  **(2.63,**  **5.26)** | **4.13**  **(1.39, 12.22)** | **4.35**  **(2.04,**  **9.30)** | **3.82**  **(1.58,**  **9.23)** | **0.984** |
| **ELISpot-NMO**  **GMR at 2-4 weeks post-boost and pre-boost (95%CI)** | **0.94**  **(0.60,**  **1.49)** | **0.89**  **(0.50,**  **1.60)** | **1.24**  **(0.46,**  **3.38)** | **1.00**  **(0.47,**  **2.11)** | **1.10**  **(0.35,**  **3.47)** | **0.947** |

Note: * *p* ≤ 0.05

Abbreviation: GM: geometric mean, SFU/10^6^ cells: spot forming units per million cells, GMR: geometric mean ratio
