## Supplementary Table S7 for "Immunogenicity and reactogenicity of mRNA COVID-19 vaccine booster administered by intradermal or intramuscular route in Thai Older adults"

**Supplementary Table S7.** Adverse events among older people aged >65 years after the booster (3rd dose) vaccination (post-boost) by type and route of booster vaccine administration

| **Adverse events** |  | **Type and route of booster vaccine administration** | | | | **p-value** |
| --- | --- | --- | --- | --- | --- | --- |
|  | **Total** | **mRNA-1273**  **(ID)** | **mRNA-1273**  **(IM)** | **BNT162b2 (ID)** | **BNT162b2 (IM)** |  |
|  | (n = 210) | (n = 35) | (n = 35) | (n = 70) | (n = 70) |  |
| **Local reaction, n (%)**  Mild, n (%)  Moderate, n (%) | 136 (64.76)  87 (41.43)  49 (23.33) | 28 (80.00)  18 (51.43)  10 (28.57) | 25 (71.43)  11 (31.43)  14(40.00) | 38 (54.29)  31 (44.29)  7(10.00) | 45 (64.29)  27 (38.57)  18 (25.71) | 0.017* |
| **Injection site reaction, n (%)**  Mild, n (%)  Moderate, n (%) | 136 (64.76)  87 (41.43)  49 (23.33) | 28 (80.00)  18 (51.43)  10 (28.57) | 25 (71.43)  11 (31.43)  14(40.00) | 38 (54.29)  31 (44.29)  7(10.00) | 45 (64.29)  27 (38.57)  18 (25.71) | 0.017* |
| **Systemic reaction, n (%)**  Mild, n (%)  Moderate, n (%) | 107 (51.43)  66 (31.43)  42 (20.00) | 14 (40.00)  8 (22.86)  6 (17.14) | 18 (51.44)  9 (25.17)  9 (25.17) | 35 (50.00)  25 (35.71)  10 (14.29) | 41 (58.57)  24 (34.29)  17 (24.29) | 0.067 |
| **Myalgia, n (%)**  Mild, n (%)  Moderate, n (%) | 70 (33.33)  42 (20.00)  28 (13.33) | 9 (25.71)  4 (11.43)  5 (14.29) | 13 (37.14)  7 (20.00)  6 (17.14) | 17 (24.28)  11 (15.71)  6 (8.57) | 31 (44.28)  20 (28.57)  11 (15.71) | 0.137 |
| **Fatigue, n (%)**  Mild, n (%)  Moderate, n (%) | 39 (18.57)  28 (13.33)  11 (5.24) | 5 (14.29)  4 (11.43)  1 (2.86) | 3 (8.57)  2 (5.71)  1 (2.86) | 14 (20.00)  12 (17.14)  2 (2.86) | 17 (24.29)  10 (14.29)  7 (10.00) | 0.100 |
| **Headache, n (%)**  Mild, n (%)  Moderate, n (%) | 35 (16.67)  15 (7.14)  20 (9.52) | 6 (17.14)  2 (5.71)  4 (11.43) | 5 (14.29)  1 (2.86)  4 (11.43) | 8 (11.43)  5 (7.14)  3 (4.29) | 16 (22.86)  7 (10.00)  9 (12.86) | 0.331 |
| **Fever, n (%)**  Mild, n (%)  Moderate, n (%) | 7 (3.33)  5 (2.38)  2 (0.95) | 0 (0.00)  0 (0.00)  0 (0.00) | 2 (5.72)  1 (2.86)  1 (2.86) | 4 (5.71)  3 (4.29)  1 (1.43) | 1 (1.43)  1 (1.43)  0 (0.00) | 0.231 |
| **Flu symptoms, n (%)**  Mild, n (%)  Moderate, n (%) | 2 (0.01)  0 (0.00)  2 (0.01) | 0 (0.00)  0 (0.00)  0 (0.00) | 0 (0.00)  0 (0.00)  0 (0.00) | 0 (0.00)  0 (0.00)  0 (0.00) | 2 (0.86)  0 (0.00)  2 (0.86) | 0.129 |
| **Diarrhea, n (%)**  Mild, n (%)  Moderate, n (%) | 15 (7.14)  10 (4.76)  5 (2.38) | 3 (8.57)  2 (5.71)  1 (2.86) | 0 (0.00)  0 (0.00)  0 (0.00) | 6 (8.57)  5 (7.14)  1 (1.43) | 6 (8.58)  3 (4.29)  3 (4.29) | 0.217 |
| **Nausea, n (%)**  Mild, n (%)  Moderate, n (%) | 9 (4.29)  6 (2.86)  3 (1.43) | 2 (5.72)  1 (2.86)  1 (2.86) | 1 (2.86)  1 (2.86)  0 (0.00) | 2 (2.86)  1 (1.43)  1 (1.43) | 4 (5.72)  3 (4.29)  1 (1.43) | 0.556 |
| **Rash, n (%)**  Mild, n (%)  Moderate, n (%) | 38 (18.10)  27 (12.86)  11 (5.24) | 10 (28.57)  7 (20.00)  3 (8.57) | 3 (8.57)  2 (5.71)  1 (2.86) | 22 (31.43)  16 (22.86)  6 (8.57) | 3 (4.29)  2 (2.86)  1 (1.43) | <0.001* |
| **Vomit, n (%)**  Mild, n (%)  Moderate, n (%) | 8 (3.81)  6 (2.86)  2 (0.95) | 0 (0.00)  0 (0.00)  0 (0.00) | 0 (0.00)  0 (0.00)  0 (0.00) | 4 (5.71)  3 (4.29)  1 (1.43) | 4 (5.71)  3 (4.29)  1 (1.43) | 0.268 |

Note: * *p* ≤ 0.05
