## Supplementary Table S8 for "Immunogenicity and reactogenicity of mRNA COVID-19 vaccine booster administered by intradermal or intramuscular route in Thai Older adults"

**Supplementary Table S8.** Adverse events among older people aged 65-79 years and ≥ 80 years after the booster (3rd dose) vaccination (post-boost) by type and route of booster vaccine administration

| **Adverse events** | **Type and route of booster vaccine administration** | | | | | | | | | | **p-value** |
| --- | --- | --- | --- | --- | --- | --- | --- | --- | --- | --- | --- |
|  | **Total**  (n = 210) | | **mRNA-1273 (ID)**  (n = 35) | | **mRNA-1273 (IM)**  (n = 35) | | **BNT162b2 (ID)**  (n = 70) | | **BNT162b2 (IM)**  (n = 70) | |  |
|  | **≥ 80**  **Years**  (n = 92) | **65-79 years**  (n = 118) | **≥ 80**  **Years**  (n = 11) | **65-79 years**  (n = 24) | **≥ 80 years**  (n = 12) | **65-79 years**  (n= 23) | **≥ 80**  **Years**  (n = 35) | **65-79 years**  (n = 35) | **≥ 80**  **Years**  (n = 34) | **65-79 years**  (n = 36) |  |
| **Local reaction, n (%)**  Mild, n (%)  Moderate, n (%) | 54 (58.70)  35 (38.04)  19 (20.66) | 83 (70.34)  52 (44.07)  31 (26.27) | 10 (90.91)  3 (27.27)  7 (63.64) | 17 (70.83)  15 (62.50)  2 (83.33) | 8 (66.67)  5 (41.67)  3 (25.00) | 17 (73.91)  6 (26.09)  11 (47.82) | 15 (42.86)  14 (40.00)  1 (2.86) | 23 (65.71)  17 (48.57)  6 (17.14) | 21 (61.77)  13 (38.24)  8 (23.53) | 24 (66.67)  14 (38.89)  10 (27.78) | 0.013* |
| **Injection site reaction, n (%)**  Mild, n (%)  Moderate, n (%) | 54 (58.70)  35 (38.04)  19 (20.66) | 83 (70.34)  52 (44.07)  31 (26.27) | 10 (90.91)  3 (27.27)  7 (63.64) | 17 (70.83)  15 (62.50)  2 (83.33) | 8 (66.67)  5 (41.67)  3 (25.00) | 17 (73.91)  6 (26.09)  11 (47.82) | 15 (42.86)  14 (40.00)  1 (2.86) | 23 (65.71)  17 (48.57)  6 (17.14) | 21 (61.77)  13 (38.24)  8 (23.53) | 24 (66.67)  14 (38.89)  10 (27.78) | 0.013* |
| **Systemic reaction, n (%)**  Mild, n (%)  Moderate, n (%) | 46 (50.00)  31 (33.70)  15 (16.30) | 62 (52.54)  35 (29.66)  27 (22.88) | 4 (33.36)  0 (0.00)  4 (33.36) | 10 (41.67)  8 (33.33)  2 (8.34) | 6 (50.00)  5 (41.67)  1 (8.33) | 12 (52.17)  4 (17.39)  8 (34.78) | 19 (54.29)  16 (45.71)  3 (8.58) | 16 (45.71)  9 (25.71)  7 (20.00) | 17 (50.00)  10 (29.41)  7 (20.59) | 24 (66.67)  14 (38.89)  10 (27.78) | 0.024* |
| **Myalgia, n (%)**  Mild, n (%)  Moderate, n (%) | 28 (30.43)  18 (19.57)  10 (10.86) | 42 (35.90)  24 (20.35)  18 (15.25) | 4 (33.36)  0 (0.00)  4 (33.36) | 5 (20.84)  4 (16.67)  1 (4.17) | 3 (25.00)  3 (25.00)  0 (0.00) | 10 (43.48)  4 (17.39)  6 (26.09) | 8 (24.24)  6 (18.18)  2 (6.06) | 9 (25.71)  5 (14.29)  4 (11.42) | 13 (38.24)  9 (26.47)  4 (11.76) | 18 (50.00)  11 (30.56)  7 (19.44) | 0.179 |
| **Fatigue, n (%)**  Mild, n (%)  Moderate, n (%) | 18 (19.57)  16 (17.39)  2 (2.18) | 22 (18.64)  13 (11.02)  9 (7.62) | 1 (9.09)  1 (9.09)  0 (0.00) | 4 (16.67)  3 (12.50)  1 (4.17) | 0 (0.00)  0 (0.00)  0 (0.00) | 3 (13.04)  2 (8.70)  1 (4.34) | 10 (43.48)  10 (17.39)  0 (26.09) | 5 (14.28)  3 (8.57)  2 (5.71) | 7 (20.59)  5 (14.71)  2 (5.88) | 10 (27.78)  5 (13.89)  5 (13.89) | 0.295 |
| **Headache, n (%)**  Mild, n (%)  Moderate, n (%) | 14 (15.22)  8 (8.70)  6 (6.52) | 21 (17.80)  7 (5.94)  14 (11.86) | 2 (18.18)  0 (0.00)  2 (18.18) | 4 (16.67)  2 (8.33)  2 (8.34) | 1 (8.33)  1 (8.33)  0 (0.00) | 4 (17.39)  0 (0.00)  4 (17.39) | 4 (11.43)  4 (11.43)  0 (0.00) | 4 (11.43)  1 (2.86)  3 (8.57) | 7 (20.58)  3 (8.82)  4 (11.76) | 9 (25.71)  4 (11.42)  5 (14.29) | 0.349 |
| **Fever, n (%)**  Mild, n (%)  Moderate, n (%) | 2 (2.17)  2 (2.17)  0 (0.00) | 5 (4.23)  3 (2.54)  2 (1.69) | 0 (0.00)  0 (0.00)  0 (0.00) | 0 (0.00)  0 (0.00)  0 (0.00) | 0 (0.00)  0 (0.00)  0 (0.00) | 2 (8.70)  1 (4.35)  1 (4.35) | 2 (5.71)  2 (5.71)  0 (0.00) | 2 (5.72)  1 (2.86)  1 (2.86) | 0 (0.00)  0 (0.00)  0 (0.00) | 1 (2.78)  1 (2.78)  0 (0.00) | 0.622 |
| **Flu symptoms, n (%)**  Mild, n (%)  Moderate, n (%) | 0 (0.00)  0 (0.00)  0 (0.00) | 2 (1.69)  2 (1.69)  0 (0.00) | 0 (0.00)  0 (0.00)  0 (0.00) | 0 (0.00)  0 (0.00)  0 (0.00) | 0 (0.00)  0 (0.00)  0 (0.00) | 0 (0.00)  0 (0.00)  0 (0.00) | 0 (0.00)  0 (0.00)  0 (0.00) | 0 (0.00)  0 (0.00)  0 (0.00) | 0 (0.00)  0 (0.00)  0 (0.00) | 2 (5.56)  2 (5.56)  0 (0.00) | 0.246 |
| **Diarrhea, n (%)**  Mild, n (%)  Moderate, n (%) | 4 (4.35)  3 (3.26)  1 (1.09) | 11 (9.32)  7 (5.93)  4 (3.39) | 0 (0.00)  0 (0.00)  0 (0.00) | 3 (12.50)  2 (8.33)  1 (4.17) | 0 (0.00)  0 (0.00)  0 (0.00) | 0 (0.00)  0 (0.00)  0 (0.00) | 3 (8.57)  3 (8.57)  0 (0.00) | 3 (8.57)  2 (5.71)  1 (2.86) | 1 (2.94)  0 (0.00)  1 (2.94) | 5 (13.89)  3 (8.33)  2 (5.56) | 0.542 |
| **Nausea, n (%)**  Mild, n (%)  Moderate, n (%) | 1 (1.09)  1 (1.09)  0 (0.00) | 8 (6.78)  5 (4.24)  3 (2.54) | 0 (0.00)  0 (0.00)  0 (0.00) | 2 (8.34)  1 (4.17)  1 (4.17) | 0 (0.00)  0 (0.00)  0 (0.00) | 1 (4.35)  1 (4.35)  0 (0.00) | 1 (2.86)  1 (2.86)  0 (0.00) | 1 (2.86)  0 (0.00)  1 (2.86) | 0 (0.00)  0 (0.00)  0 (0.00) | 4 (11.11)  3 (8.33)  1 (2.78) | 0.632 |
| **Rash, n (%)**  Mild, n (%)  Moderate, n (%)3 | 14 (15.22)  10 (10.87)  4 (4.35) | 24 (20.34)  17 (14.41)  7 (5.93) | 3 (27.27)  1 (9.09)  2 (18.18) | 7 (29.17)  6 (25.00)  1 (4.17) | 0 (0.00)  0 (0.00)  0 (0.00) | 3 (13.04)  2 (8.70)  1 (4.34) | 10 (28.57)  8 (22.86)  2 (5.71) | 12 (34.29)  8 (22.86)  4 (11.43) | 1 (2.94)  1 (2.94)  0 (0.00) | 2 (5.72)  1 (2.86)  1 (2.86) | 0.005* |
| **Vomit, n (%)**  Mild, n (%)  Moderate, n (%) | 4 (4.35)  4 (4.35)  0 (0.00) | 4 (3.38)  2 (1.69)  2 (1.69) | 0 (0.00)  0 (0.00)  0 (0.00) | 0 (0.00)  0 (0.00)  0 (0.00) | 0 (0.00)  0 (0.00)  0 (0.00) | 0 (0.00)  0 (0.00)  0 (0.00) | 3 (8.57)  3 (8.57)  0 (0.00) | 1 (2.86)  0 (0.00)  1 (2.86) | 1 (2.94)  1 (2.94)  0 (0.00) | 3 (8.33)  2 (5.56)  1 (2.77) | 0.253 |

Note: * *p* ≤ 0.05
