## Supplementary figures and images for "Immunogenicity and reactogenicity of mRNA COVID-19 vaccine booster administered by intradermal or intramuscular route in Thai Older adults"

### Supplementary Figure S1

**Supplementary Figure S1.** Consort diagram of the study.

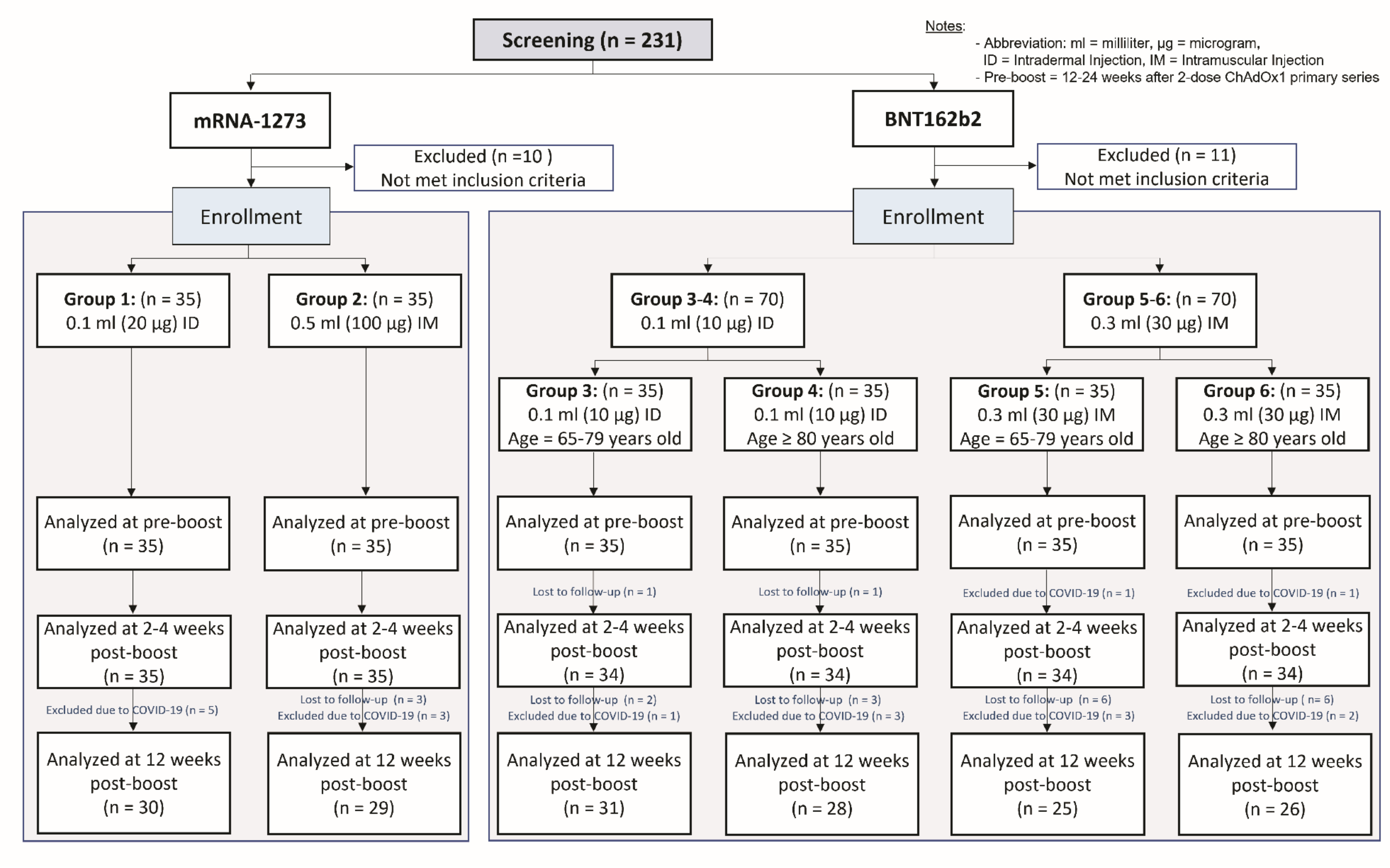
